# Single-cell Transcriptome Analysis Indicates New Potential Regulation Mechanism of ACE2 and NPs signaling among heart failure patients infected with SARS-CoV-2

**DOI:** 10.1101/2020.04.30.20081257

**Authors:** Dachun Xu, Mengqiu Ma, Yanhua Xu, Yang Su, Sang-Bing Ong, Xingdong Hu, Min Chai, Maojun Zhao, Hong Li, Yingjie Chen, Xiaojiang Xu

**Author notes:** These authors contributed equally (Dachun Xu, Mengqiu Ma, Yanhua Xu, Yang Su). **Subject Terms:** ACE/Angiotensin Receptors/Renin Angiotensin System. **Address correspondence to:** Correspondence: Dr. Xiaojiang Xu, National Institute of Environmental Health Sciences, National Institutes of Health, 111 TW Alexander Dr, Research Triangle Park, NC, 27709, USA.

## Abstract

**Background:** COVID-19 patients with comorbidities such as hypertension or heart failure (HF) are associated with poor clinical outcomes. Angiotensin-converting enzyme 2 (ACE2), the critical enzyme for SARS-CoV-2 infection, is broadly expressed in many organs including heart. However, the cellular distribution of ACE2 in the human heart, particularly the failing heart is unknown.

**Methods:** We analyzed single-cell RNA sequencing (scRNA-seq) data in both normal and failing hearts, and characterized the ACE2 gene expression profile in various cell subsets, especially in cardiomyocyte subsets, as well as its interaction with gene networks relating to various defense and immune responses at the single cell level.

**Results:** The results demonstrated that ACE2 is present in cardiomyocytes (CMs), endothelial cells, fibroblasts and smooth muscle cells in the heart, while the number of ACE2-postive (ACE2+) CMs and ACE2 gene expression in these CMs are significantly increased in the failing hearts. Interestingly, both brain natriuretic peptides (BNP) and atrial natriuretic peptide (ANP) are significantly up-regulated in the ACE2+ CMs. Further analysis shows that ANP, BNP and ACE2 may form a negative feedback loop with a group of genes associated with the development of heart failure. To our surprise, we found that genes related to virus entry, virus replication and suppression of interferon-gamma (IFN-γ) signaling are all up-regulated in CMs in failing hearts, and the increases were significantly higher in ACE2+ CMs as compared with ACE2 negative (ACE2-) CMs, suggesting that these ACE2+ CMs may be more vulnerable to virus infection. Since ACE2 expression is correlated with BNP expression, we further performed retrospective analysis of the plasma BNP levels and clinic outcome of 91 COVID-19 patients from a single-center. Patients with higher plasma BNP were associated with significantly higher mortality rate and expression levels of inflammatory and infective markers such as procalcitonin and C-reactive protein.

**Conclusion:** In the failing heart, the upregulation of ACE2 and virus infection associated genes, as well as the increased expression of ANP and BNP could facilitate SARS-CoV-2 virus entry and replication in these vulnerable cardiomyocyte subsets. These findings may advance our understanding of the underlying molecular mechanisms of myocarditis associated with COVID-19.

## Introduction

Novel coronavirus disease 2019 (COVID-19) is an infectious disease caused by severe acute respiratory syndrome coronavirus 2 (SARS-CoV-2). SARS-CoV-2 virus enters human cells via binding its surface “spike” to bind angiotensin-converting enzyme 2 (ACE2)^1^. SARS-CoV-2 has spread worldwide and was classified as a pandemic in 2020. As of May 2020, more than four million cases of COVID-19 and more than 276,000 deaths have been reported worldwide^2^. In addition to the severe lung infection, the SARS-CoV-2 also causes myocarditis, cardiac dysfunction, and heart failure (HF)^1, 3, 4^. On the other hand, COVID-19 patients with any pre-existing conditions, such as hypertension, coronary heart disease, and cardiac injury, have worse clinical outcomes than those without these comorbidity ^5, 6^. Thus, statistical results indicate a vicious cycle between SARS-CoV-2 infection and cardiac dysfunction or HF ^5^.

ACE2 is the critical enzyme degrading the pro-inflammatory angiotensin-II to the anti-inflammatory Ang 1-7 ^7,8^. Unfortunately, ACE2 also facilitates SARS-CoV-2 infection in host cells. ACE2 is highly expressed in the nose, kidney, intestine, colon, brain, endothelium, testis, and heart^9-14^. A recent study from Zou *et al*. reported that ~7% cardiomyocytes (CMs) express ACE2 in normal human cardiac tissues^11^, suggesting that some CMs can be directly infected by SARS-CoV-2. However, ACE2 gene expression in different cardiomyocyte subsets, as well as its dynamic changes in failing human hearts at the single cell level, are totally unknown.

Since ACE2 plays an important role in SARS-CoV-2 infection and cardiac function, it is critically important to understand its distribution and the biological changes associated with alterations in its expression in normal and failing hearts. Therefore, we investigated the ACE2 gene expression profiles by analyzing the single-cell RNA sequencing (scRNA-seq) dataset derived from both normal and failing human hearts^15^. Interestingly, we found that ACE2 was selectively expressed in some of ventricular and atrial CMs, vascular endothelial cells, fibroblasts, smooth muscle cells and immune cells in both normal and failing hearts, and its expression was further increased in several cell subsets in the failing hearts. Importantly, we found that brain natriuretic peptide (BNP) and atrial natriuretic peptide (ANP) transcripts are co-upregulated in ACE2-postive (ACE2+) CMs. BNP, ANP, and ACE2 may form a feedback loop associated with the RAAS (rein-angiotensin-aldosterone-system)/Ang II signaling pathway. Furthermore, ACE2 expression was also associated with the dynamic changes of a group of genes related to viral infection and acquired immunity. Since there is a positive correlation between the expressions of BNP and ACE2, we further analyzed the clinic outcome, inflammation markers, and blood BNP levels in COVID-19 patients retrospectively. Together, these findings provide important insights to advance our understanding of the interplays between ACE2, viral infection and inflammation, as well as cardiac injury and failure.

## Materials and Methods

### Study design and Participants

This retrospective, single-center study included 91 patients with laboratory-confirmed COVID-19 admitted to Ezhou Central Hospital, Ezhou, China from January 25, 2020 and March 30, 2020. PCR-Fluorescence probing based kit (Novel Coronavirus (2019-nCoV) Nucleic Acid Diagnostic Kit, Sansure Biotech, China) was used to extract nucleic acids from clinical samples and to detect the ORF1ab gene (nCovORF1ab) and the N gene (nCoV-NP) according to the manufacturer’s instructions. SARS-CoV-2 infection was laboratory-confirmed if the nCovORF1ab and nCoV-NP tests were both positive results. The study protocol was approved by the ethics committee of Shanghai Tenth People’s Hospital, Tongji University School of Medicine (Shanghai, China). Patient informed consent was waived by each ethics committee due to the COVID-19 pandemic.

COVID-19 was diagnosed by meeting at least one of these two criteria: (i) chest computerized tomography (CT) manifestations of viral pneumonia; and/or (ii) reverse transcription-polymerase chain reaction (RT-PCR) according to the New Coronavirus Pneumonia Prevention and Control Program (5th edition) published by the National Health Commission of China (New Coronavirus Pneumonia Prevention and Control Program. 2020) and WHO interim guidance^16^. We used the following inclusion and exclusion criteria to determine the study cohort. The inclusion criteria were confirmed COVID19, valid BNP level and aged above 18 years. The exclusion criteria were incomplete medical records, pregnancy, acute myocardial infarction, acute pulmonary embolism, acute stroke, HIV infection, and preexisting organ failure (chronic cirrhosis, chronic renal failure, or severe congestive heart failure and end-stage cancer).

### Data Collection

The demographic characteristics and clinical data (comorbidities, laboratory findings, and outcomes) for participants during hospitalization were collected from electronic medical records. Cardiac biomarkers measured on admission were collected, including Troponin I (TNI), creatine kinase-MB (CK-MB), and BNP. All data were independently reviewed and entered into the computer database by three analysts. Since the echocardiography data were unavailable for most patients, patients were categorized according to the BNP level. Acute HF was defined as a blood BNP level ≥100 pg/ml. The clinical outcomes (i.e., discharges and mortality) were monitored up to 30 days.

## Statistical Analysis

Descriptive statistics were obtained for all study variables. Continuous data were expressed as mean (standard deviation (SD)) or median (interquartile [IQR]) values. Categorical data were expressed as proportions. All continuous variables were compared using the t-test or the Mann-Whitney U-test if appropriate. In contrast, categorical variables were analyzed for the study outcome by Fisher exact test or χ^2^ test. The Pearson correlation coefficient and Spearman rank correlation coefficient were used for linear correlation analysis. Survival analysis between patients with BNP<100 pg/mL and ≥100 pg/mL was conducted by the Kaplan-Meier estimate with p-value generated by the log-rank test. Data were analyzed using SPSS version 25.0 (IBM Corp) or Graphpad Prism 8.0.1 (GraphPad Software, San Diego, CA). For all the statistical analyses, 2-sided p<0.05 was considered significant.

## scRNA-seq analysis

### Data Sources

Adult human heart scRNA-seq datasets were obtained from Gene Expression Omnibus (GEO) under accession codes GSE109816 and GSE121893. Briefly, samples from twelve healthy donors and samples from six patients with HF were collected at the time of heart transplantation. The range of donor ages was 21-52 year, with a median age of 45.5 years.

### Sequencing data processing

The processed read count matrix was retrieved from existing sources based on previously published data as specified explicitly in the reference. Briefly, Raw reads were processed using the Perl pipeline script supplied by Takara.

### Single-cell clustering and identified cell types

The processed read count matrix was imported into R (Version 3.6.2) and converted to a Seurat object using the Seurat R package (Version 3.1.2). Cells that had over 75% UMIs derived from the mitochondrial genome were discarded. For the remaining cells, gene expression matrices were normalized to total cellular read count using the negative binomial regression method implemented in Seurat *SCTransform* function. Cell-cycle scores were calculated using Seurat *CellCycleScoring* function. The Seurat *RunPCA* functions were performed to calculate principal components (PCs). We further corrected the batch effect using Harmony because batch effects among the human heart samples were observed. The *RunUMAP* function with default setting was applied to visualize the first 35 Harmony aligned coordinates. The *FindClusters* function with resolution=0.2 parameter was carried out to cluster cells into different groups. Canonical marker genes were applied to annotate cell clusters into known biological cell types. Monocle 3^17^ as used to perform trajectory and pseudotime analysis.

### Identification of differential expression genes (DEGs)

To identify DEG between two groups, we applied the Seurat *FindMarkers* function with the default parameter of method “MAST” and cells ID from each defined group (e.g. ACE2+ cells versus ACE2 negative (ACE2-) cells in CM1) as input.

### Gene function analysis

GSEA (Version 4.03) was used to perform gene ontology (GO) term and pathway enrichment analysis with the Molecular Signatures Database (MSigDB, C2 and C5, Version 7.01).

## Results

### Integrated analysis of normal and HF conditions at single-cell resolution

To detect the discrepancy between normal and HF patients, we utilized the scRNA-seq data by Wang *et al*^15^. Briefly, twelve control samples were collected from healthy donor hearts (hereinafter called normals). Samples from six HF patients were collected at the time of heart transplantation. 9767 out of 9994 cells from normals and 4219 out of 4221 cells from patients passed standard quality control and were retained for subsequent analyses. On average, 1649 and 1904 genes were detected in individual cells from normals and patients, respectively. We performed uniform manifold approximation and projection (UMAP) and clustering analysis and grouped the entire population into nine subsets (Fig. 1A). Dot plot showed the expression of known markers for nine clusters, which included: 1) endothelial cells (Cluster 1, PECAM1 and VWF); 2) fibroblasts (Cluster 5, LUM and DCN); 3) smooth muscle cells (Cluster 3, MYH11); 4) NK-T/ monocytes (Cluster 6, CD3G and CD163); 5) granulocytes (Cluster 9, HP, ITLN1); 6) CM2 and 3 subsets (Clusters 2/4/8, MYH6 and NPPA); 7) CM1 and 4 subsets (Clusters 0 and 7, MYH7 and MYL2) (Fig. 1B). Then, UMAP for individual sample was separately plotted side by side and exhibited the differential distribution of subsets between normal and HF patients. As shown in Fig. 1C, all nine subsets were detected in both normal and patient groups. However, the percentage of CM1 was dramatically decreased (39.65% in normals versus 6.71% in HF, p<0.0001), while the percentage of CM4 was significantly increased (0.03% in normals versus 8.70% in HF, p<0.0001) in HF samples. In addition, the percentages of CM2 (17.70% in normals versus 18.68% in HF, p>0.05) and CM3 (8.27% in normals versus 8.53% in HF, p>0.05) were significantly decreased in HF samples. The percentages of vascular endothelial cells (16.79% in normal versus 28.13% in HF, p<0.0001) and fibroblasts (4.53% in normal versus 8.41% in HF, p<0.0001) were also significantly increased in the failing hearts (Fig. 1D).

**Figure 1.**
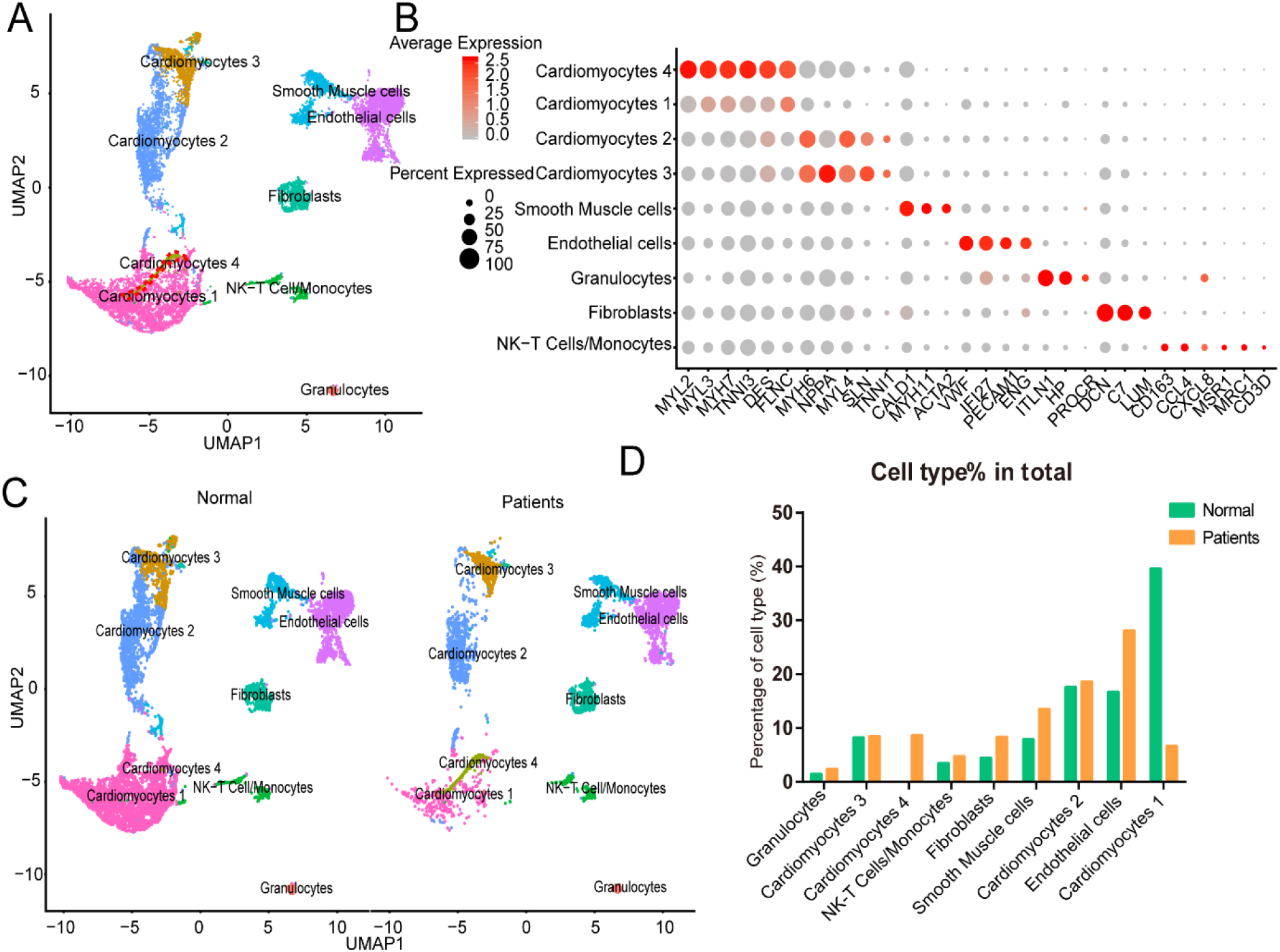
Integrated analysis of normal and heart failure (HF) conditions at single-cell resolution. **A**, Uniform manifold approximation and projection (UAMP) clustering of 14698 cells isolated from normal and heart failure patients. Each dot represents a single cell. Cell type was annotated by the expression of known marker genes. **B**, Dot plotting showing gene signature among different clusters, the shadings denotes average expression levels and the sizes of dots denote fractional expression. **C**, Split views show the 9 subsets in normal and patient group. **D**, The percentage of cell number for different cell types in normal and patient group.

For each cluster, we calculated the cluster-specific genes (marker genes). Left ventricle (LV) marker genes MYL2 and MYL3 were highly expressed in CM1 and CM4; these subsets were thus termed ventricular cardiomyocytes. Since the left atrial (LA) marker genes MYH6 and MYH7 were highly expressed in CM2 and CM3 subsets, they were termed atrial CMs^18^.

### Both CMs and Non-CMs (NCMs) show different characteristics between normal and HF patients

We compared gene expression of atrial cardiomyocytes (CM2&3) and NCMs between normal and patients. We observed that GO term viral gene expression was up-regulated in all atrial CMs and NCMs in HF (Online Fig. IA-F). These findings suggested that some CMs and NCMs in the heart may be liable to SARS-CoV-2 infection. In addition, GO results showed that genes related to the mitochondrial respiratory complexes and ATP synthesis were up-regulated, while genes related to the inflammatory response, leukocyte migration, response to interferon-gamma and defense against pathogens were downregulated in atrial cardiomyocytes in HF patients. The reduced inflammatory response may result in an increased sensitivity to SARS-CoV-2 virus infection in these atrial CMs (Online Fig. IA).

To further characterize this unusual CM4 subset observed in failing hearts, we performed trajectories analysis of the integrated clusters to show the pseudotime of CMs and NCMs. Trajectory and pseudotime results indicated that CM4 originated from CM1 (Fig. 2A), which is consistent with our speculation that CM4 may be a distinct type CM after HF. We then conducted GSEA analysis (GO and Pathway) on DEG between CM4 and CM1. Viral gene expression, as well as pathways related to influenza infection, infectious diseases, and HIV infection were upregulated in CM4 (Fig. 2B and 2C); while response to virus, defense response to virus, response to interferon gamma and innate immune response, pathway of the adaptive immune response, interferon signaling and interferon-alpha-beta-gamma signaling were significantly down-regulated in CM4 (Fig. 2D and 2E). Together, these results suggest that the CM4 subset predominantly observed in HF tissues would be more vulnerable to virus infection than the CM1 subset.

**Figure 2.**
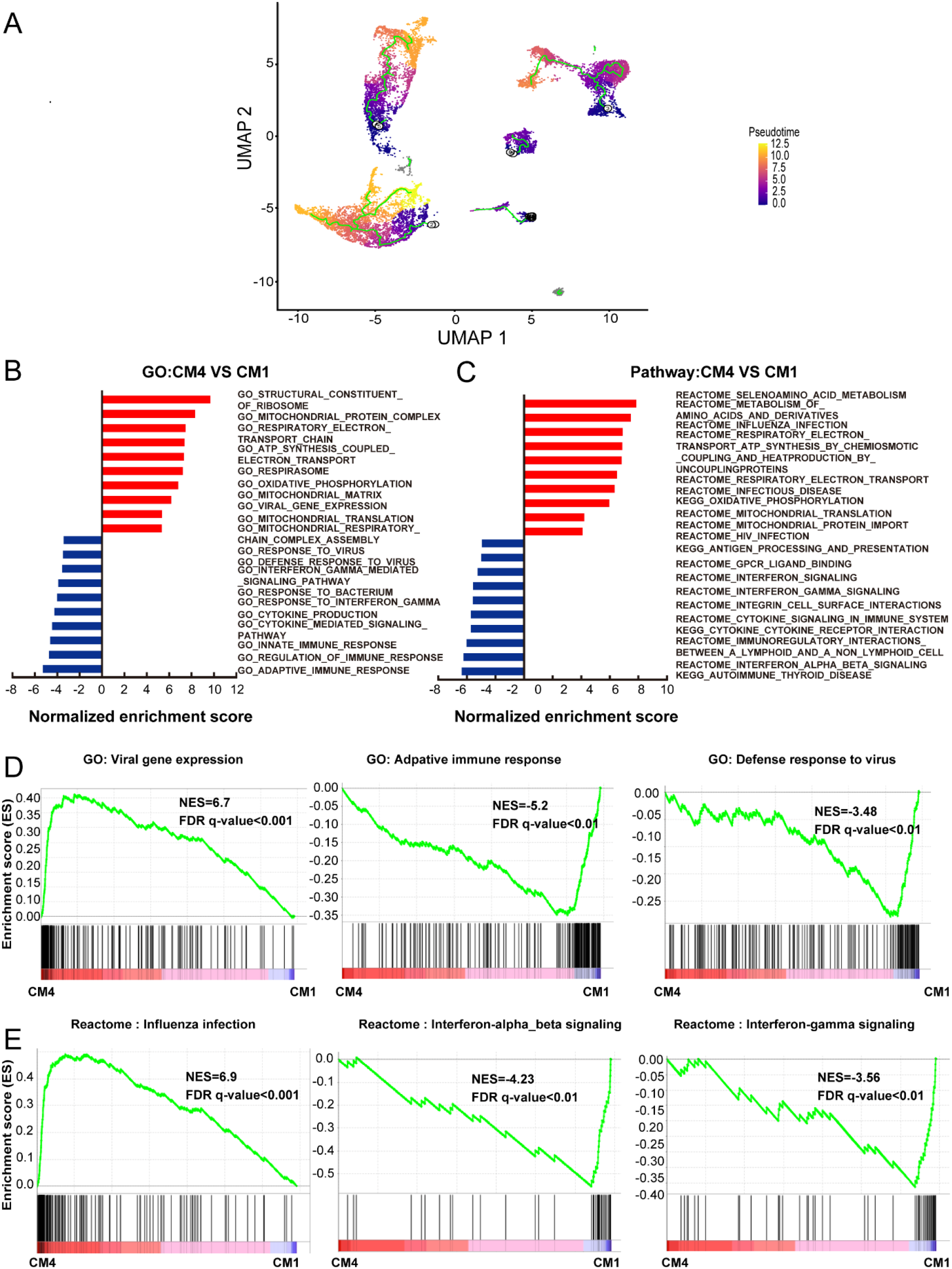
Cardiomyocytes 4 (CM4) shows different characteristics with Cardiomyocytes 1(CM1) **A**, Pseudotime analysis of the nine clusters, the color from purple to yellow denote the different developing stage, and the simultaneous principal curve indicates the pseudo-time stage. **B, C**, GSEA analysis revealed significant enrichment of GO and pathways for CM4 compared with CM1. **D**, GO enrichment showing GO terms of increased viral gene expression, decreased adaptive immune response and defense response to virus. **E**, Influenza infection signaling pathway is up-regulated, both interferon-alpha-beta signaling and interferon-gamma signaling are down-regulated.

### Both CMs and NCMs have different ACE2 expression pattern after HF

We further investigated the frequency of ACE2+ cells frequency in CMs and NCMs in normal and failing hearts. Fig. 3A showed the overall distribution of ACE2+ cells in different subsets of normal and HF samples. The frequency of ACE2+ cells increased significantly in three of four CMs in HF patients, especially in CM1 and CM4. The percentages of ACE2+ cells increased from 5.55% to 34.98% in CM1 subset (p<0.0001), and increased from 0% to 7.01% (p<0.0001) in CM4 subset. The percentage of ACE2+ cells in CM3 subset significantly increased from 6.19% to 13.16% (p<0.0001), while its frequency in CM2 subset did not change significantly (5.55% in normal versus 5.71% in HF, p>0.05) (Fig. 3B). Moreover, the percentages of ACE2+ cells in fibroblasts (p<0.0001) and smooth muscle cells (p=0.0104) were both significantly decreased. The frequency of ACE2+ cells in NK-T Cell/Monocytes increased from 3.77% to 5.42% (p>0.05), while its percentage in granulocytes was not significantly changed (2.04% in normal versus 5.83% in HF patients, p> 0.05).

**Figure 3.**
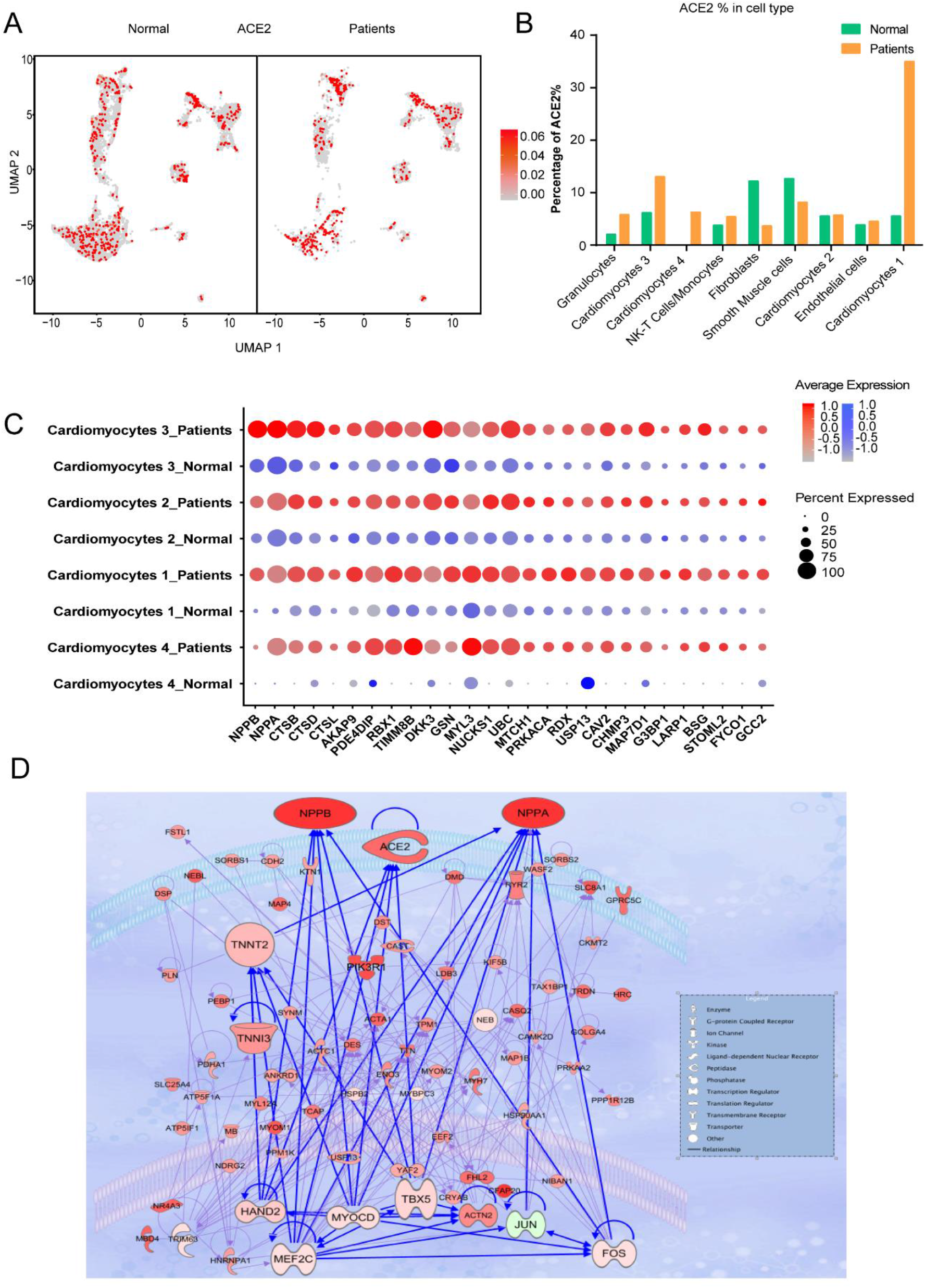
Cardiomyocytes (CMs) and Non-CMs (NCMs) have different ACE2 expression pattern. **A**, UAMP of the CMs and NCMs subsets in normal and HF patients. **B**, Frequency of ACE2+ cells in different cell types. **C**, Gene expression pattern of virus infection-related genes in different subsets of CMs during HF. **D**, Gene regulatory network of ACE2, NPPA, NPPB and TNNT1,2,3. and their upstream binding transcription factor of HAND2, MYOCD, MEF2C and TBX5.

Taken together, scRNA-seq results demonstrated that the ACE2+ CMs dramatically increased during HF, suggesting that CMs in HF patients may be more susceptible to SARS-CoV-2 virus infection than the normal subjects. In addition, ventricular myocytes had a higher percentage of ACE2+ cells than that of atrial myocytes, indicating that these cardiomyocyte subsets may have different responses to SARS-CoV-2 infection.

### Virus infection-related genes are upregulated in CMs in HF patients

We then focused on gene expression dynamics of the SARS-CoV-2 entry receptor ACE2. To further examine the potential role of ACE2+ cells in the myocardium infected by SARS-CoV-2, we separated each cardiomyocyte subset into two sub-groups according to the expression of ACE2 (ACE2+ and ACE2-) and called DEGs between these two groups.

One of the most interesting findings was that *NPPB* (the gene coding BNP) and *NPPA* (the gene coding ANP) were the top two upregulated genes in ACE2+ cells as compared to ACE2-cells, and the increases were over 1.8 fold for both genes. Previous studies reported that ACE2, ANP, BNP, TnT and TnI could make a feedback loop to preserve ejection fraction in HF patients^19-22^. Interestingly, most of the ejection fraction preservation genes were significantly upregulated during HF, especially in ACE2+ CMs cells (Fig. 3C). We used the top 100 DEGs of ACE2+ and ACE2- in CM1,4 to build a gene regulatory network (GRN) using IPA (Ingenuity Pathway Analysis, QIAGEN, CA, USA). GRN showed that *ACE2, NPPA, NPPB, AGT, TNNT1, TNNT2* and *TNNT3* were well connected and shared the same upstream binding transcription factors HAND2, MYOCD, MEF2C, TBX5 which are the well-known transcription factors that can control the reprogramming of fibroblasts into CMs(Fig. 3D) ^23, 24^ The above findings further suggest that these cardiac ejection fraction preservation genes may affect SARS-CoV-2-induced cardiomyocyte infection and injury of cardiac myocytes, as their expression is correlated with that of ACE2.

We further studied the expression dynamics of *ACE2*, *NPPA* and *NPPB* in CMs and NCMs in normal and HF patients. Both *NPPB* and *NPPA* were co-expressed with *ACE2* and significantly up-regulated in CMs in HF samples (Fig. 4A, 4B), but *NPPB* and *NPPA* showed different expression patterns. Specifically, *NPPA* was expressed only in CM2, 3 and NCMs, but it was not expressed in CM1 and CM4 subsets in normal heart. *NPPA* was expressed in all CMs and NCMs, and its expression was significantly upregulated in all cardiomyocyte subsets after HF (Fig. 4B). *NPPB* was only expressed in CM2 and CM3 subsets in normal heart, and its expression was significantly upregulated in CM2, CM3, and CM1 subsets after HF (Fig. 4A). Pro-ANP and pro-BNP can be processed by corin and intracellular endoprotease furin in in vitro experiments to form active ANP and BNP, respectively^25, 26^. We found that in HF patients, corin expression increased significantly in CMs while the change in furin was insignificant (Online Fig. IIA), which is consistent with the observation that furin activity, but not its concentration, increased ^27^. Importantly, at the S1/S2 boundary of SARS-CoV-2, a furin cleavage site has been identified, which can enhance the binding of spike protein and host cells^28^. It was reported that Polypeptide N-Acetylgalactosaminyltransferase, such as B3GALNT1, GALNT1 can mediate the glycosylation of proBNP and increase proBNP secretion in human cardiac during HF^29^. Both B3GALNT1 and GALNT1 transcription increased in HF patients (Online Fig. IIB). We then assessed other virus infection-related genes, which are involved in virus entry (BSG, CAV2, CHMP3, CHMP5, STOML2), cysteine proteases cathepsins (CSTB, CSTD, CSTL), virus replication (AKAP9, RDX, 16 MTCH1) and suppression of IFN-γ signaling (LARP1, RBX1 and TIMM8B) (Fig. 4C-F). Genes contributing to virus entry (Fig. 4C,4D, Online Fig. IIC, IID), virus replication (Fig. 4F) and suppression of IFN-γ signaling (Fig. 4E) were all up-regulated in CMs in failing hearts.

It was reported that SARS-CoV-2 enters host cells through the binding of its spike protein with ACE2 and subsequent S protein priming by host cell protease TMPRSS2^31,30^. To our surprise, we barely detected any expression of TMPRSS2 in both normal and HF samples (Online Fig. IIE). Since it is reported that in the absence of cell surface protease TMPRSS2, SARS-CoV can achieve cell entry via an endosomal pathway in which it can be activated by other proteases such as cathepsin L^30^, we further investigated gene expression dynamics of the endosomal cysteine proteases, cathepsins and found out that CTSB, CTSD and CTSL were up-regulated significantly in CMs during HF (Fig. 4D). We also detected that the expression levels of some inflammatory cytokines were increased in several subsets in the HF patients, such as CXCL8 which was significantly increased in the subset of granulocytes and NK-T cell/Monocytes as well as IL-32 which was increased in the subsets of NK-T cell/Monocytes and endothelial cells, respectively (Online Fig. IIF).Thus, we speculate that SARS-CoV-2 may use the ACE2-CTSB/L axis for cell entry in cardiac tissues. Together, these findings suggest that failing hearts might be more vulnerable to SARS-CoV-2 infection.

**Figure 4.**
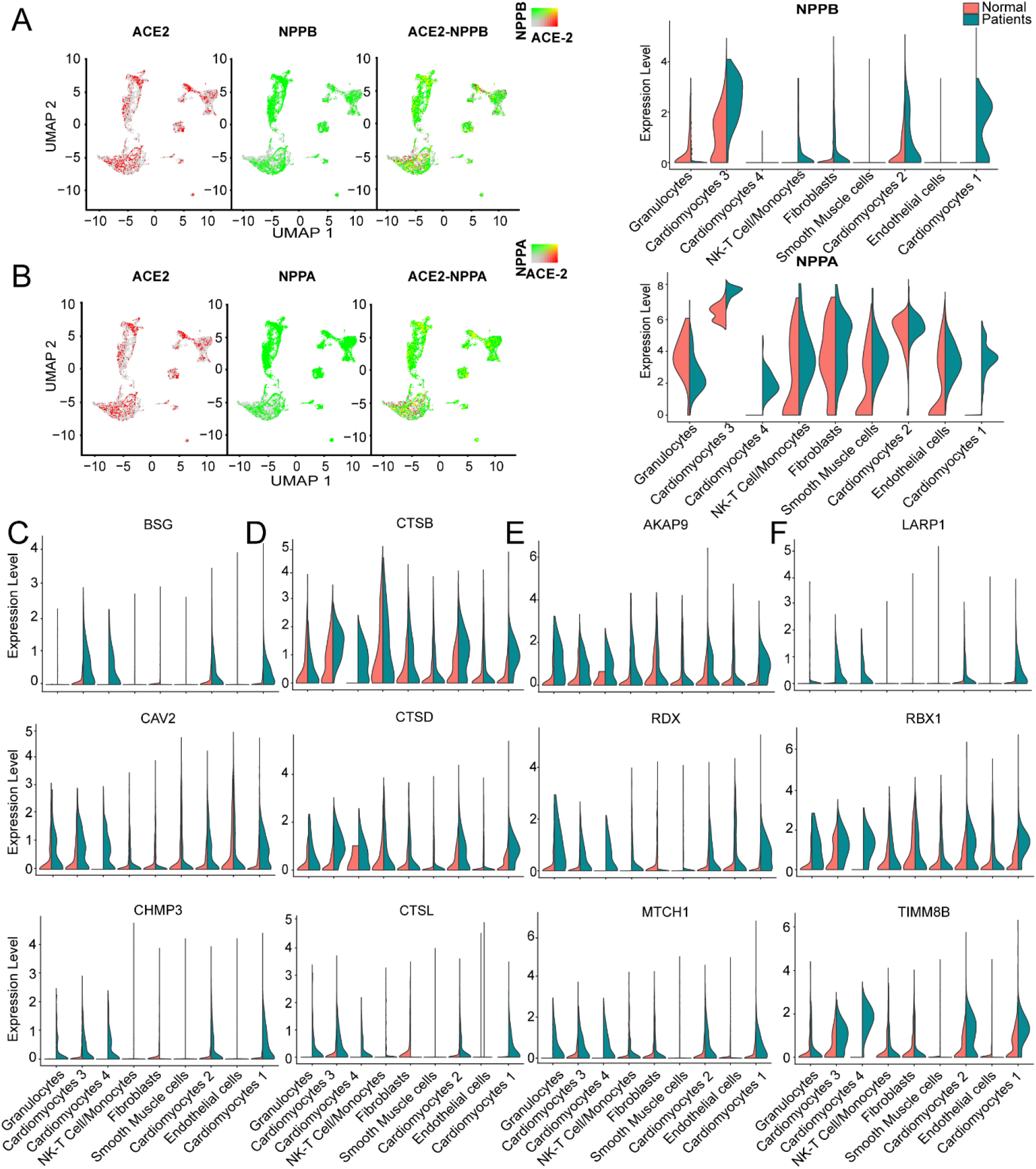
Virus related genes are upregulated in heart failure (HF) patients compared with normal. **A**, Expression level of ACE2 (red dots), NPPB (green dots) in different clusters, overlapping is shown in the right panel, and the co-expression is shown in yellow dots. Violin plots of the distribution of NPPB between normal and HF patients in different subsets. **B**, Expression level of ACE2 (red dot), NPPA (green dot) in different subsets, overlapping is shown in the right panel, and the co-expression is shown in yellow dots. Violin plots of the distribution of NPPA between normal and HF patients in different subsets. **C**, Violin plots of the distribution of genes (from top to bottom BSG, CAV2, CHMP3) related to viral infection. **D**, Violin plots of the gene expression pattern of CST B/L. **E**, Violin plots of the distribution of genes (from top to bottom AKAP9, RDX, MTCH1) related to IFN-γ signaling pathway. **F**, Violin plots of the distribution of genes (from top to bottom LARP1, RBX1 and TIMM8B) on viral replication.

Thrombosis is commonly observed in severe COVID-19 patients^32^. Tissue factor (TF/CD142) activation causes thrombus formation on atherosclerotic plaques coded by F3^33^. We investigated the expression dynamics of genes related to blood clotting. F3 was co-expressed with ACE2 and significantly up-regulated in CM3 and CM1 during HF (Online Fig. IIG, IIH), suggesting that increased F3 and ACE2 may contribute to the increased risk of thrombosis in HF patients.

### Characteristics of ACE2-positive ventricular and atrial CMs, and NCMs

We further conducted GSEA analysis on DEGs of cells between ACE2+ and ACE2- in CM1 and CM4 (Fig. 5A, Online Fig. IIIA). GO terms associated with energy consumption (Fig. 5A), energy derivation by oxidation (Fig. 5C), and pathway influenza infection (Online Fig. IIIC) and infectious disease (Online Fig. IIIA) were positively enriched in ACE2+ cells. In contrast, GO terms associated with interferon gamma-mediated signaling pathway, defense response to virus and interferon-alpha_beta signaling and interferon signaling were negatively enriched in ACE2+ cells (Fig. 5C, Online Fig. IIIC).

**Figure 5.**
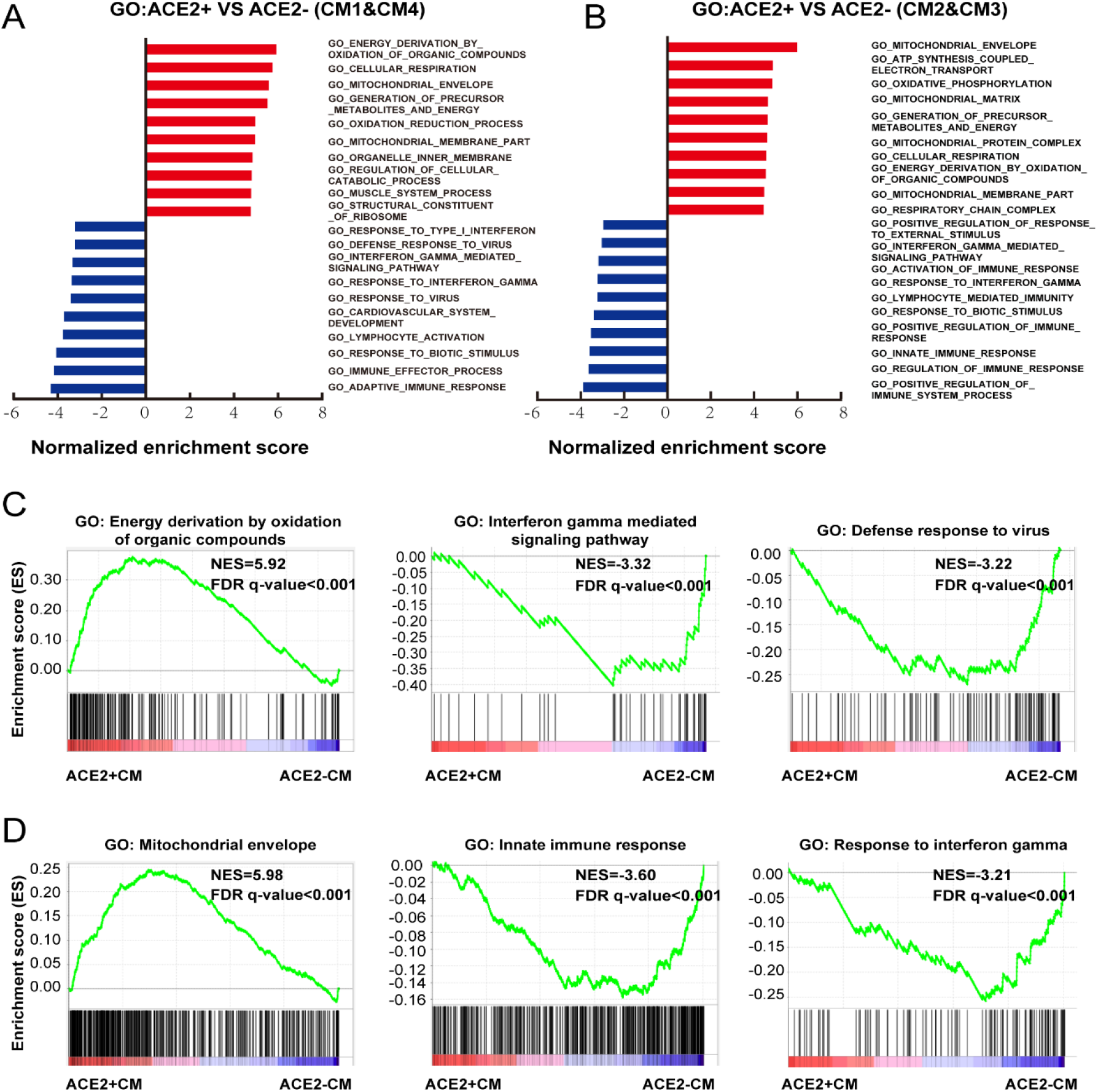
Characteristics of ACE2+ ventricular and atrial myocytes. **A**, GO analysis revealed significant enrichment of biological pathways for ACE2+ compared with ACE2- in ventricular myocytes. **B**, GO analysis revealed significant enrichment of biological pathways for ACE2+ compared with ACE2- in atrial myocytes. **C**, GO plots showing GO terms of increased energy derivation by oxidation of organic compounds (left), decreased interferon gamma mediated signaling pathway (median) and down-regulated defense response to virus (right). **D**, GO enrichment plots showing GO terms of increased mitochondrial envelope (left), decreased innate immune response (median) and down-regulated innate immune response (right). The NES and false discovery rate (FDR) were showed in panel.

We also performed GSEA analysis on DEGs of cells between ACE2+ and ACE2- in CM2 and CM3 (Fig. 5B, Online Fig. IIIB). GO terms associated with energy consumption, mitochondrial envelope, ATP synthesis coupled electron transport, oxidative phosphorylation, pathway cardiac muscle contraction, and respiratory electron transport were positively enriched (Fig. 5B, 5C, Online Fig. IIIB, IIIC). GO terms and pathways associated with innate immune response, response to interferon gamma, interferon gamma signaling and interferon-alpha_beta signaling were negatively enriched, which are consistent with the observation in ventricular CMs (Fig. 5D, Online Fig. IIID).

Moreover, we also identified DEGs between ACE2+ NCMs and ACE2-NCMs and performed GSEA analysis on them (Online Fig. IV). Interestingly, pathways associated with infectious disease were positively enriched in NCMs, except for NK-T Cells/Monocytes. GO terms associated with mitochondrial matrix and ATP synthesis were positively enriched in smooth muscle cells, NK-T Cells/Monocytes and fibroblasts, which is consistent with the observation at CMs. GO term associated with muscle structure and function (Online Fig. IVA, IVE) and leukocyte mediated immunity were negatively enriched in ACE2+ cells of smooth muscle cells, fibroblasts, and endothelial cells (Online Fig. IVB). GO term associated with viral expression is positively enriched in ACE2+ granulocytes, while GO term associated with immunocyte mediated immunity is negatively enriched in ACE2+ granulocytes and ACE2+ NK-T Cells/Monocytes. These findings suggest an impaired cellular immunological response in HF patients, which may increase their vulnerability to various pathogens (Online Fig. IVC, IVD).

### Clinical Characteristics of COVID-19 patients

The median age of these 91 COVID-19 patients was 66 years (range, [27-89]). 46 patients (50.5%) have elevated BNP (≥100 pg/mL). HF patients have increased BNP plasma concentrations which are generally corelated with the degree of cardiac dysfunction. Thus, BNP is often used as a biochemical marker for HF^34^. Patients with a higher BNP were older (median age, 71 [IQR 44-89] vs. 62 [27-79], p<0.0001) (Table 1). Compared with the lower BNP group, patients in the higher BNP group have significantly higher levels of white blood cells (p<0.0001) and neutrophils (p<0.0001), although significantly lower number of lymphocytes (p<0.0001) (Table 1). The high BNP group has significant increased procalcitonin (p<0.0001) and C-reactive protein (p<0.0001) as compared with the low BNP group (Table 1). The high BNP group also showed imbalanced electrolyte levels and aberrant coagulation profiles as compared with the low BNP group. Furthermore, more severe organ dysfunction was observed in the high BNP group, including worse liver function indicated by higher aspartate transaminase (p<0.03), direct bilirubin (p<0.005), and lactate dehydrogenase (p<0.0001) (Table 1)..

**Table 1.** Comparison of COVID-19 patient characteristics between BNP groups.

| Parameters | Total<br>(N=91) | BNP<100<br>(N=45) | BNP≥100<br>(N=46) | p value |
| --- | --- | --- | --- | --- |
| Age, yrs, median [min, max] | 66 [27-89] | 62 [27-79] | 71 [44-89] | <0.0001* |
| Male, n (%) | 54 (59.3) | 23 (51.1) | 31 (67.4) | 0.11 |
| **Complete blood cell count, 10<sup>9</sup>/L** |  |  |  |  |
| White blood cell, median (IQR) | 7.99 (4.59-13.31) | 6.28 (4.04-8.38) | 13.05 (6.76-18.13) | <0.0001* |
| Neutrophil, median (IQR) | 6.6 (3.43-12.32) | 4.29 (2.74-6.67) | 11.88 (4.83-16.93) | <0.0001* |
| Lymphocyte, median (IQR) | 0.71 (0.38-1.09) | 0.98 (0.62-1.47) | 0.50 (0.27-0.78) | <0.0001* |
| **Liver and renal function** |  |  |  |  |
| Alanine transaminase, U/L, median (IQR) | 30.0 (18.5-52.5) | 27.0 (19.0-49.0) | 32.0 (18.0-64.0) | 0.5783 |
| Aspartate transaminase, U/L, median (IQR) | 37.0 (23.0-55.0) | 30.0 (20.0-51.0) | 41.5 (29.0-64.0) | 0.0299* |
| TBIL, µmol/L, median (IQR) | 14.1 (9.5-21.7) | 11.8 (9.1-17.8) | 15.2 (10.1-24.4) | 0.1216 |
| Direct bilirubin, µmol/L, median (IQR) | 5.3 (3.4-9.8) | 4.1 (3.0-6.5) | 6.6 (4.0-13.2) | 0.0047* |
| Lactate dehydrogenase, U/L, median (IQR) | 315.0 (179.5-470.5) | 185.0 (154.0-352.0) | 407.0 (288.0-599.0) | <0.0001* |
| eGFR, mL/(min*1.73m<sup>2</sup>), mean±SD | 105.6±47.0 | 121.1±41.6 | 86.5±44.5 | 0.0003* |
| Blood urea nitrogen, mmol/L, median (IQR) | 5.7 (3.9-11.1) | 4.5 (3.2-5.8) | 9.0 (5.2-15.9) | <0.0001* |
| Uric acid, µmol/L, median (IQR) | 234.0 (183.5-305.5) | 235.0 (184.0-305.0) | 230.5 (182.0-310.0) | 0.9494 |
| **Cardiac biomarker** |  |  |  |  |
| Troponin-I, ng/mL, median (IQR) | 0.01 (0.01-0.06) | 0.01 (0.01-0.01) | 0.05 (0.03-0.25) | <0.0001* |
| **Electrolytes** |  |  |  |  |
| Potassium, mmol/L, median(IQR) | 4.04 (3.64-4.40) | 3.87 (3.56-4.27) | 4.19 (3.64-4.70) | 0.0354* |
| Sodium, mmol/L, median (IQR) | 139.0 (136.0-142.0) | 139.0 (135.0-141.0) | 139.0 (136.0-145.0) | 0.2992 |
| Chloride, mmol/L, median (IQR) | 102.0 (98.5-106.0) | 103.0 (100.0-106.0) | 101.5 (98.0-106.0) | 0.6473 |
| Calcium, mmol/L, mean±SD | 2.03±0.18 | 2.09±0.16 | 1.97±0.18 | 0.0018* |
| **Coagulation profiles** |  |  |  |  |
| Prothrombin time, s, median (IQR) | 13.4 (12.4-14.9) | 13.0 (12.0-13.8) | 13.9 (12.8-16.7) | 0.0030* |
| APTT, s, median (IQR) | 35.5 (31.8-39.8) | 35.1 (32.4-38.9) | 35.5 (30.7-42.5) | 0.6165 |
| Fibrinogen, g/L, median (IQR) | 3.39 (2.31-4.77) | 3.39 (2.31-5.09) | 3.40 (2.35-5.87) | 0.8567 |
| D-dimer, µg/mL, median (IQR) | 2.03 (1.22-1.00) | 1.37 (0.83-1.99) | 6.96 (3.25-24.20) | <0.0001* |
| **Inflammatory biomarkers** |  |  |  |  |
| Procalcitonin, ng/mL, median(IQR) | 0.45 (0.12-1.12) | 0.23 (0.04-0.49) | 1.01 (0.39-3.51) | <0.0001* |
| hsCRP, mg/L, median (IQR) | 13.80 (5.74-20.50) | 6.09 (1.52-15.86) | 18.00 (13.45-21.50) | <0.0001* |
| **Blood gas analysis** |  |  |  |  |
| PaO<sub>2</sub>, mmHg, median (IQR) | 71.0 (57.8-92.0) | 78.5 (57.5-104.5) | 68,5 (56.5-86.0) | 0.4867 |
| PaCO<sub>2</sub>, mmHg, median (IQR) | 41.0 (34.0-48.8) | 39.5 (33.5-43.5) | 42.5 (34.0-57.9) | 0.1589 |
| Lactic acid, mmol/L, median (IQR) | 1.95 (1.40-2.40) | 1.80(1.30-2.15) | 2.00 (1.60-2.75) | 0.1634 |
| **BNP**, pg/mL, median (IQR) | 92.0 (32.5-299.5) | 34.0 (15.0-48.0) | 299.5 (180.0-548.0) | **<0.0001\*** |
| **Death**, n (%) | 32 (35.16) | 5 (11.11) | 27 (58.70) | **<0.0001\*** |

Continuous variables are presented as means ± SD if they conform to normal distribution, or median with interquartile range if not. Age is presented as median with range (minimum to maximum). Categorical variables are presented as percentage (%). \* Significant p value (<0.05). TBIL denotes total bilirubin; eGFR, estimated glomerular filtration rate (calculated by MDRD formula); APTT, activated partial thromboplastin time; hsCRP, high-sensitive C-reactive protein; BNP, brain natriuretic peptide.

The high BNP group also showed worse renal function as indicated by a reduced glomerular filtration rate (<0.0003) and increased blood urea nitrogen (p<0.0001) (Table 1). Cardiac TNI (p<0.0001) was significantly increased in the higher BNP group, suggesting more cardiac injury in these patients (Table 1). Noteworthy, the high BNP group had a higher incidence of respiratory failure (RF, 31.43%, p=0.0064) (Fig. 6A left), and a significantly increased mortality rate (58.70%, p<0.0001) (Fig. 6A middle, Table 1), and a negative correlation with the lymphocyte count (Fig. 6A right). Infective markers were positively correlated with the BNP level (Fig. 6B). Markers of coagulative disturbance and organ impairment were positively correlated with the BNP level (Fig. 6C middle and right).

**Figure 6.**
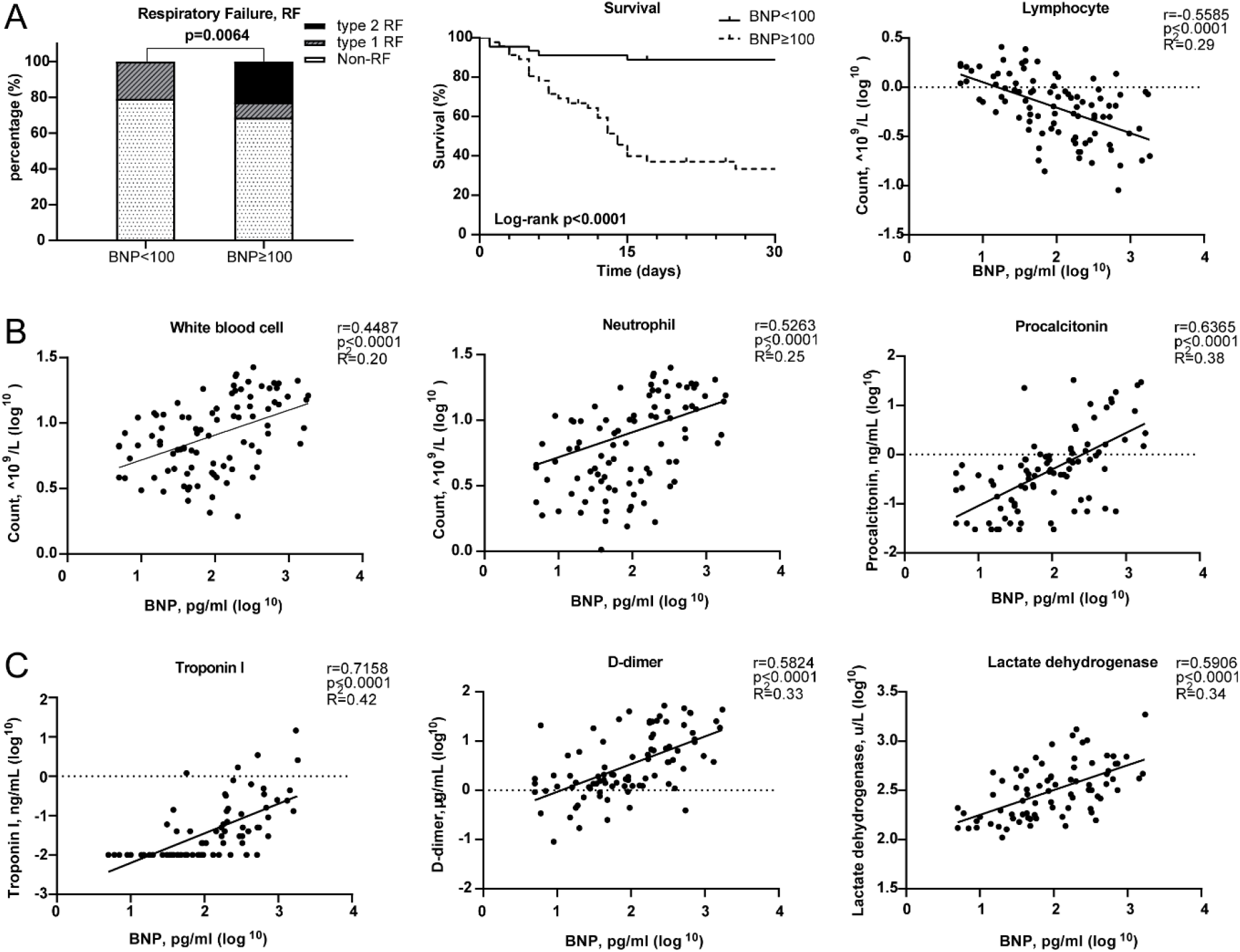
Relationships between Brain natriuretic peptide (BNP) level and clinical assessments. **A**, Measurement of disease severity. Left figure described the constitution of non-respiratory failure (RF), type 1 RF, and type 2 RF patients. Middle figure showed K-M estimation of the mortality in high BNP group. Right figure showed the significant negative correlation of lymphocyte count and BNP level. **B**, The severity of infection. From left to right, white blood cell, neutrophil and lymphocyte counts were positively correlated with BNP level. **C**, The relationship between organ impairment and BNP. Left figure depicted the strong positive relationship of cardiac injury and blood BNP level. Middle figure showed the disturbance of coagulation as BNP level increased. Right figure showed the positive correlation between lactate dehydrogenase and BNP level.

## Discussion

The present research has several major findings. First, the study systematically investigated the ACE2 expression dynamics in ventricular CMs, atrial CMs, endothelial cells, fibroblast and leukocytes in human normal and failing hearts at the single-cell level. We found that ACE2 was expressed in some, but not all, of the ventricular and atrial CMs, vascular endothelial cells, and smooth muscle cells in both normal and failing hearts. Second, we demonstrated that ACE2 expression was selectively increased in the dominant ventricular CM1 subset, an unusual ventricular CM4 subset, and the atrial CM3 subset. The expression of ACE2 transcripts was also increased in these cells. Third, we demonstrated for the first time that BNP and ANP transcripts are markedly enriched in ACE2+ CMs, while BNP, ANP, and ACE2 can form a feedback loop associated with the RAAS/Ang II signaling pathway. Fourth, we demonstrated for the first time that ACE2 expression was associated with the dynamic changes of a group of genes specific for the networks of viral infection and immunity in cardiomyocytes. Moreover, we found that compared with COVID-19 patients with a lower blood BNP, those with a higher BNP had a significantly higher mortality rate and expression levels of inflammatory and infective markers such as C-reactive protein and procalcitonin. These findings provide new insights to advance our understanding of the potentially important roles of ACE2 and the associated critical signaling pathways in regulating virus infection, immunologic responses, and associated cardiomyocyte injury in normal and failing hearts.

One of the most interesting findings is that ACE2 was not equally expressed in all of the ventricular and atrial CMs, but only expressed in ~5% normal ventricular or atrial CMs. ACE2 expression was increased to 30% in the major ventricular CM1 subset in failing hearts. ACE2 was also increased in an unusual ventricular CM4 subset and in the atrial CM3 subset but was unchanged in the atrial CM2 subset. Meanwhile, ACE2 expression was unchanged in the vascular endothelial subset but decreased in the vascular smooth muscle subset in heart failure samples. Our finding that ACE2 was expressed in normal hearts appears to contradict a previous report that pericytes (with marker genes ABCC9 and KCNJ8), but not the cardiomyocytes express ACE2 in normal hearts^35^. The discrepancy may due to the fact that the previous study used the single nucleus RNA-seq approach, which generally captures many fewer transcripts as compared with the more sensitive and comprehensive SMART-seq using whole-cell in our study. While it is difficult to fully understand the pathological role of the selective alterations of ACE2 in particular cardiomyocyte subsets in the failing hearts, since ACE2 expression is required for host cell entry by SARS-CoV-2 and other coronavirus ^36^, in the context that SARS-CoV-2 causes myocarditis and cardiac injury, it is reasonable to believe that the increased ACE2 in CMs in the failing heart could exacerbate cardiac SARS-CoV-2 infection in HF patients. The finding that ACE2 was only selectively expressed in a fraction of CMs suggest that not all of the CMs in the heart are equally vulnerable to SARS-CoV-2 injury. The selective SARS-CoV-2 infection in ACE2+ CMs could certainly cause or exacerbate cardiac injury and consequent cardiac dysfunction. Moreover, the different ACE2 expression and the potential selective injury to a group of ACE2+ atrial CMs could potentially cause or exacerbate the cardiac arrhythmias that are commonly observed in HF patients. Whether SARS-CoV-2 indeed selectively causes particular atrial and ventricular CMs certainly deserves further investigations. Moreover, if our speculation regarding the increased cardiac arrhythmia in COVID-19 patients is correct, corresponding treatment should be developed.

In addition to its important role in SARS-CoV-2 and other coronavirus infections, ACE2 plays an important role in controlling the RAAS through converting Ang I and Ang II into Ang 1–9 and Ang 1–7, respectively^28, 37^. Thus, both loss-of-function and gain-of-function approaches in experimental studies have defined a critical role for ACE2 in protecting the heart against HF, systemic and pulmonary hypertension, myocardial infarction, and diabetic cardiomyopathy^19, 38, 39^. As experimental studies support an important role for ACE2 in various cardiovascular diseases and ARDS, increasing/activating ACE2 may protect against hypertension and CVD^37, 38, 40^. Previous studies have consistently demonstrated that when both SARS-CoV and SARS-CoV-2 bind to ACE2 result in loss of ACE2 function, which is driven by endocytosis, activation of proteolytic cleavage and machining^41, 42^. If ACE2 indeed protects heart and lung function in COVID-19 patients, the ACE2 degradation by SARS-CoV-2 infection contribute to the heart and lung dysfunction in these patients. In support of the above concern, a recent study demonstrated that the plasma Ang-II level from SARS-CoV-2 infected patients was markedly elevated and the plasma Ang-II linearly correlated with the viral load and lung injury in COVID-19 patients ^2, 37^, suggesting that diminished ACE2 expression might lead to the elevation of Ang-II, and consequent activation of the AT1R axis ^43^. Indeed, a recent study demonstrated that in hospitalized COVID-19 patients with hypertension, patient’s use of ACEI/ARB was associated with lower risk of all-cause mortality compared with ACEI/ARB non-users^42^. Additional clinical and experimental studies are clearly needed to define the role of ACE2 in cardiac and lung function in COVID-19 patients, and to illustrate the detailed underlying molecular mechanism of cardiac injury and HF in COVID-19 patients, as well as the mechanism of increased mortality rate in older patients and HF patients in COVID-19 patients.

Another very interesting finding in the present study is that both BNP and ANP transcripts are markedly enriched in ACE2+ CMs, and that BNP, ANP, and ACE2 can form a feedback loop associated with the RAAS/Ang II signaling pathway. Interestingly, we found that DEGs between ACE2+ and ACE2-ventricular myocytes showed that both BNP and ANP were the top two up-regulated genes. These findings are consistent with the report that ANP and BNP play important roles in chronic HF by synergizing with the renin-angiotensin-aldosterone system (RAAS) and sympathetic nervous system (SNS)^44^. ANP and BNP are commonly used biomarkers for cardiac injury and HF. Circulating ANP and BNP can promote diuresis, natriuresis and vasodilation, which is critical for the maintenance of intravascular volume homeostasis (Fig. 7B)^20^. In addition, GRN showed that *ACE2, NPPA, NPPB, AGT*, *TNNT1, TNNT2* and *TNNT3* were well connected and shared the same upstream binding transcription factors HAND2, MYOCD, MEF2C and TBX5, which imply that *ACE2, ANP* and *BNP* might be co-regulated during HF development. However, the detailed molecular mechanisms of increased ANP and BNP in ACE2+ cells are not clear currently.

**Figure 7.**
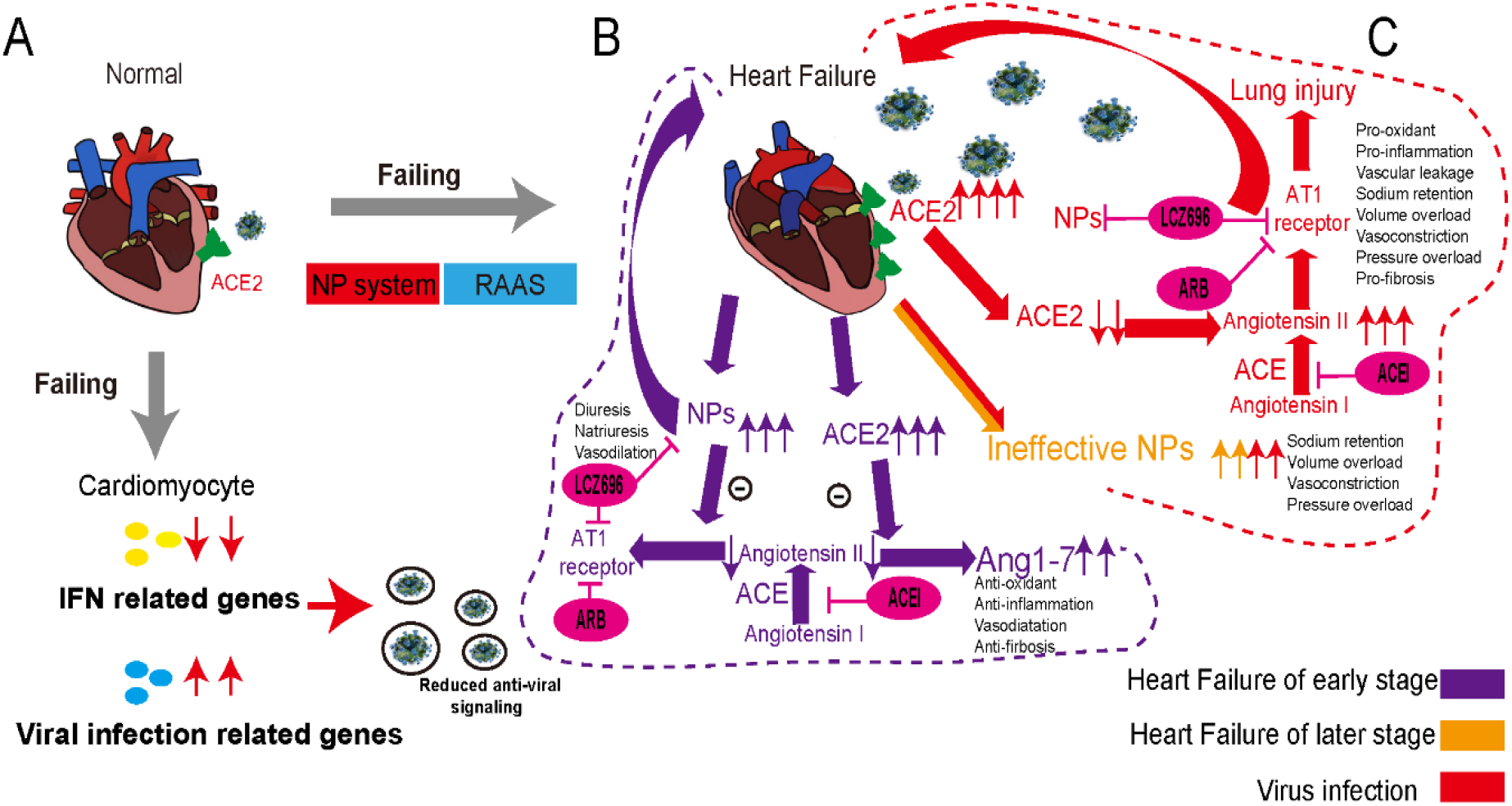
Conceptual Schematic diagram highlighting the central role of SARS-CoV-2 and the NPs, RAAS in the potentially deleterious (red) and protective (purple) effects. **A**, scRNA-seq analysis detected the down-regulated IFN related genes and up-regulated viral infection related genes during HF, which imply reduced anti-viral signaling. **B**, Schematic diagram showed the process during heart failure in the early stage and later stage noted in purple and orange, respectively. **C**, The process under virus infection was noted in red to speculate the underlying relationship for the higher susceptibility and worse prognosis in HF. Oval circles and bars indicated the potential drug and targets.

The finding that ACE2 expression was associated with the dynamic changes of a group of genes specific to viral infection and immunity in cardiomyocytes is also very interesting. In particular, we compared DEGs between failing CMs and normal CMs, as well as DEG between ACE2+ CMs with ACE-CMs. Interestingly, GSEA analysis (GO and pathway) of DEGs for these two types of comparisons achieved similar results, in which GO term/Pathway associated with viral infection and viral gene expression were positively enriched in ACE2+ cells, while defense against the virus, secretion of IFN and activation of the immune system were negatively enriched in ACE2+ CMs (Fig. 2B, 2C, 6A, 6B). These results suggest that patients with heart dysfunction or HF may have a higher susceptibility to the infection of SARS-CoV-2 and other viruses in general (Fig. 7A). Thus, the impact of ACE2 on SARS-CoV-2 in cardiac tissues could be two-fold - facilitating the virus entry to CMs and the consequent cardiomyocyte injury, and attenuating the overall virus defense capacity in cardiomyocytes. Therefore, increased ACE2 in CMs in the failing heart would certainly make these CMs more vulnerable to SARS-CoV-2.

### Limitations

First, the experimental approach used in the current study could not determine how ACE2 and NPs are synergistically regulated during HF. However, the finding could certainly encourage further investigation of the crosstalk among ACE2, ANP and BNP, as well as identifying the common transcriptional factors for these genes. Second, the novel findings presented in this study are limited at the transcriptional level. While the posttranscriptional regulation exerts critical roles in regulating the biological function for many proteins, the scRNA-seq data have clearly provided new insights to advance our understanding of the molecular mechanisms for various clinical diseases. Finally, while the present study certainly advances our understanding of ACE2 in the failing heart, the precise role of cardiac ACE2 in regulating HF patients could not be defined. Further experimental and clinical studies regarding ACE2 are warranted.

## Data Availability

All data is available online;
scRNA-seq datasets can be accessed from Gene Expression Omnibus (GEO) under accession codes GSE109816 and GSE121893

## ACKNOWLEDGMENTS

Conception and design: XX, DX. scRNA-seq data collection and analysis: XX. Provision of study materials or patients: XH, MC, MZ. Collection and assembly of clinical data: XH, MC, MZ. Results interpretation and manuscript writing: XX, MM, YX, DX, YS, YC, SBO, HL. Final approval of manuscript: XX, DX, YC, HL, SBO, YS, MM, YX, XH, MC, MZ. We thank Dr. Kenneth E. Weir and Dr. Xinli Hu for editing the manuscript. Also, we also want to thank Yuxi Sun and Teng Ma for helpful discussion.

## SOURCES OF FUNDING

This study was supported by grants from National Natural Science Foundation of China (No. 81770391 to Dachun Xu)

## DISCLOSURES

There are no conflicts of interest to declare.

## SUPPLEMENTAL MATERIALS

Supplementary material is available at Circulation research online.

Online Figures I - IV

## Nonstandard Abbreviations and Acronyms

ACE: Angiotensin-converting enzyme
ACEI: Angiotensin-converting enzyme inhibitor
ALT: Alanine transaminase
ANP: Atrial natriuretic peptide
Ang II: Angiotensin II
Ang1-7: Angiotensin1-7
ARB: Angiotensin II receptor blocker
ARDS: Acute respiratory distress syndrome
AST: Aspartate transaminase
BNP: Brain natriuretic peptide
BUN: Blood urea nitrogen
CAD: Coronary heart disease
CK-MB: Creatine kinase-MB
CMs: Cardiomyocytes
COVID-19: Coronavirus disease 2019
CRP: C-reactive protein
D-BIL: Direct bilirubin
DEG: Differential expression genes
GO: Gene ontology
GRN: Gene regulatory network
HF: Heart Failure
HR: Hazard ratio
HTN: Hypertension
IQR: Interquartile range
IRDs: Incidence rate differences
LA: Left atrium
LDL-c: Low density lipoprotein cholesterol
LDH: Lactate dehydrogenase
LV: Left ventricle
MSigDB: Molecular Signatures Database
NPs: Natriuretic Peptides
NCMs: Non-CMs
NT-proBNP: N-Terminal pro-brain natriuretic peptide
OR: Odds ratio
PCT: Procalcitonin
PT: Prothrombin time
RAAS: Renin-angiotensin-aldosterone system
SARS: Severe acute respiratory syndrome
SARS-COV: Severe acute respiratory syndrome coronavirus
scRNA-seq: Single-cell RNA sequencing
SNS: Sympathetic nervous system
TNI: Troponin I
TNT: Troponin T
UMAP: Uniform manifold approximation and projection
WHO: World Health Organization

---

**Online Figure I.**
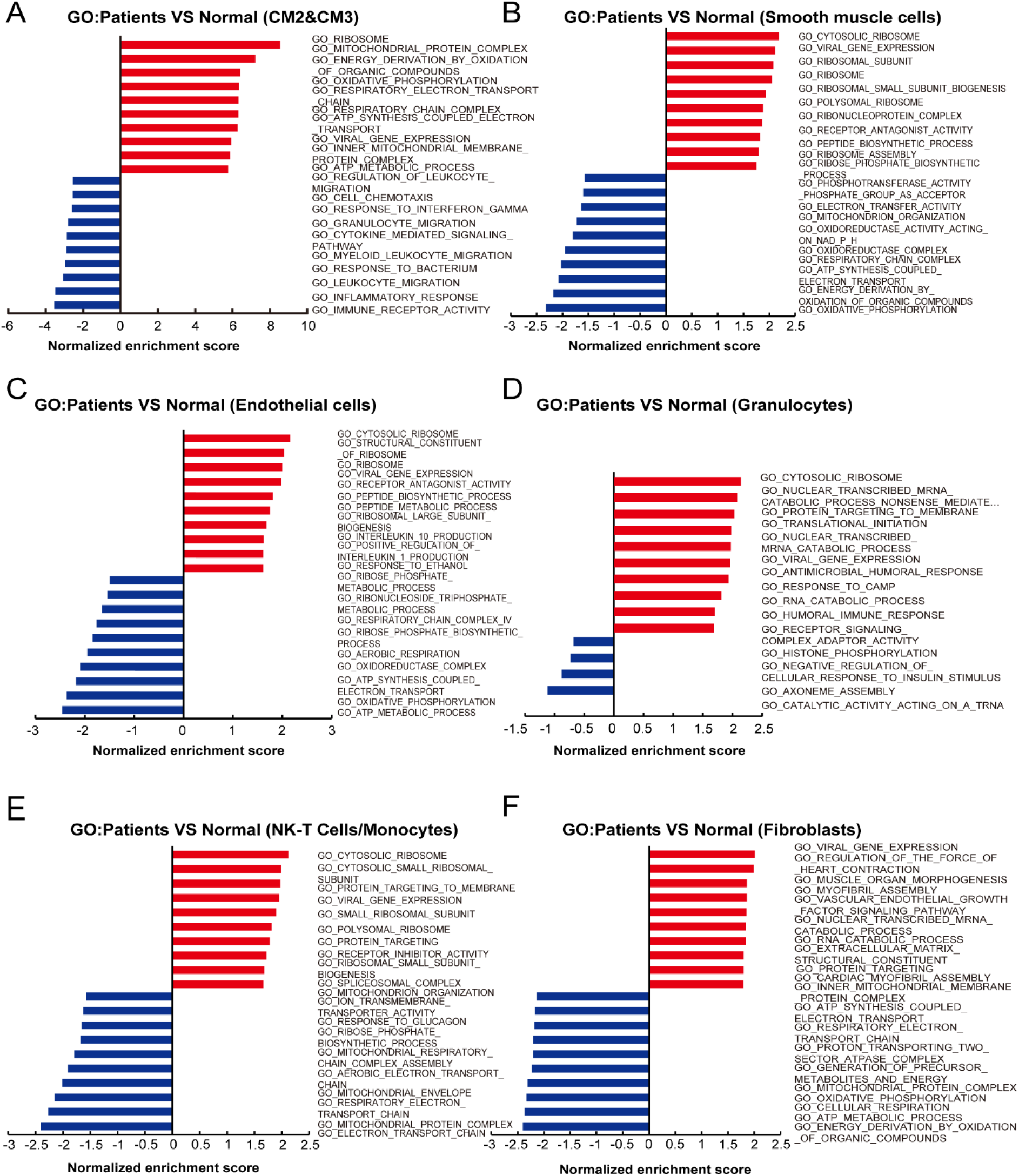
Enrichment of biological pathways in different subsets for heart failure (HF) patients compared with normals (related to Figure 2) **A-F**, GO analysis revealed significant enrichment of biological pathways for HF patients compared with normals in different subsets. **A**, Cardiomyocyte 2(CM2) and Cardiomyocyte 3(CM3) subsets. **B**, Smooth muscle cells. **C**, endothelial cells. **D**, Granulocytes. **E**, NK-T cells/Monocytes. **F**, Fibroblasts

**Online Figure II.**
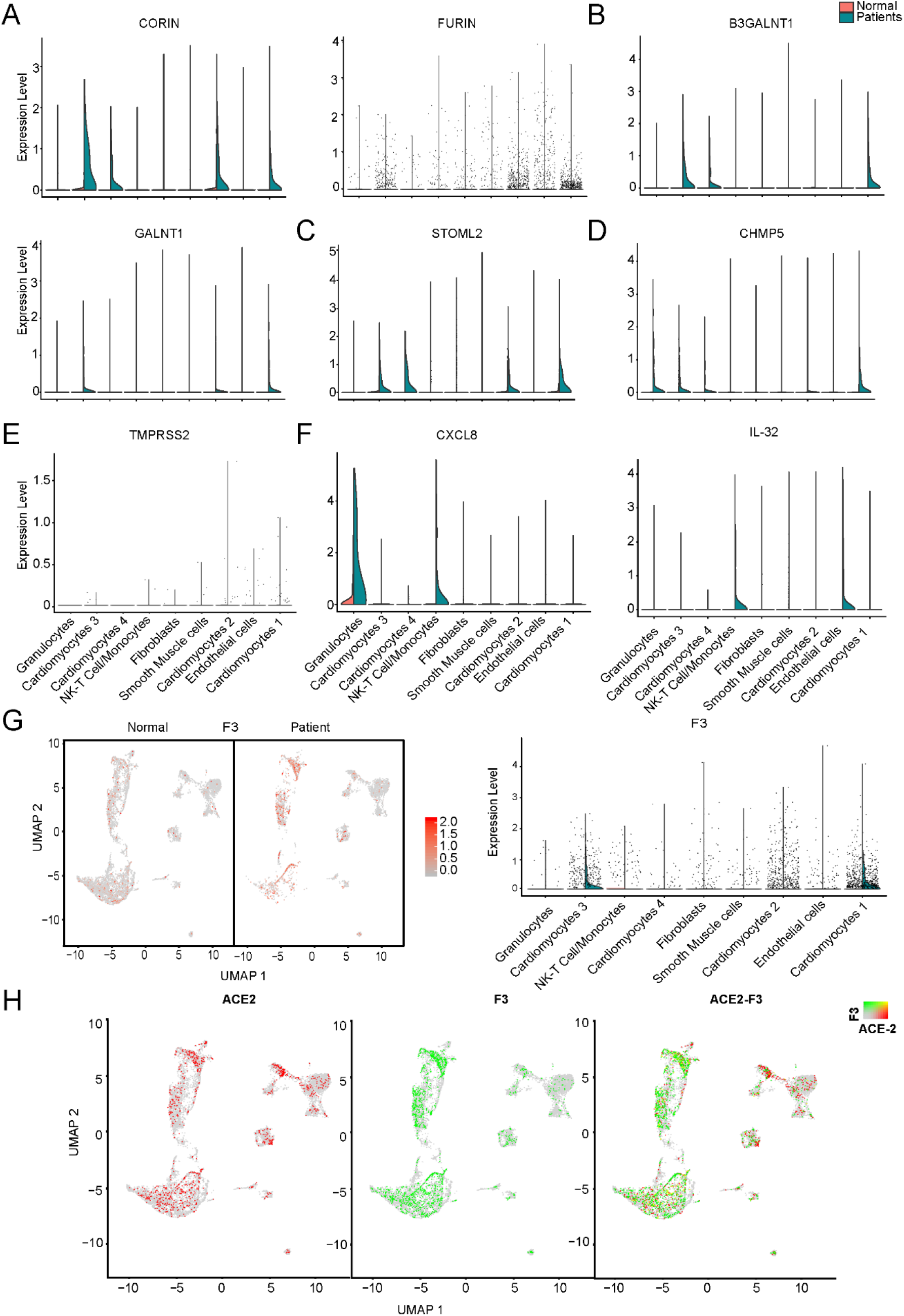
Distribution of virus-related genes for heart failure (HF) patients compared with normals (related to Figure 4) **A**, Violin plots of the distribution of STOML2 related to the biogenesis and activity of mitochondria. **B**, Violin plots of the distribution of CHMP5 related to virus infection. **C**, Violin plots of the distribution of TMPRSS2 related to viral entry. **D**, UMAP of F3 in normal and HF patients (left) and violin plots of the distribution of F3 (right).**E**, Expression level of ACE2 (red dots), F3 (green dots) in different subsets, overlapping is shown in the right panel, and the co-expression is shown in yellow dots.

**Online Figure III.**
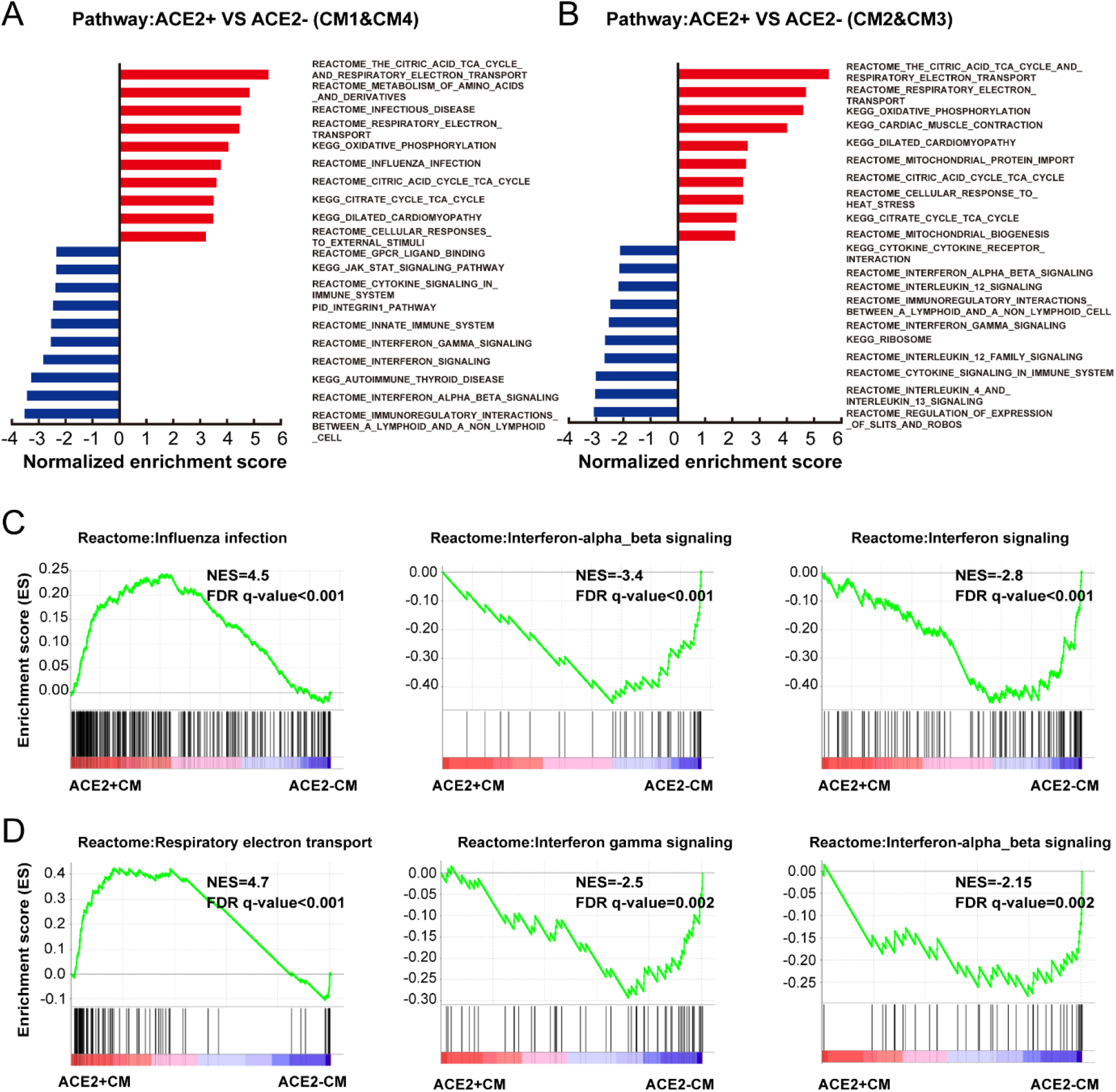
Characteristics of ACE2+ ventricular and atrial myocytes (related to Figure 5) **A**, Pathway analysis revealed the significant enrichment of biological pathways for ACE2+ compared with ACE2- in ventricular myocytes. **B**, Pathway analysis revealed the significant enrichment of biological pathways for ACE2+ compared with ACE2- in atrial myocytes. **C**, Reactome analysis showing the up-regulated influenza infection and down-regulated interferon-alpha_beta signaling and interferon signaling for ACE2+ compared with ACE2- cells in ventricular myocytes. **D**, Reactome analysis showing the up-regulated respiratory electron transport and down-regulated interferon gamma, interferon-alpha_beta signaling for ACE2+ compared with ACE2- in atrial myocytes. The NES and false discovery rate (FDR) were showed in panel.

**Online Figure IV.**
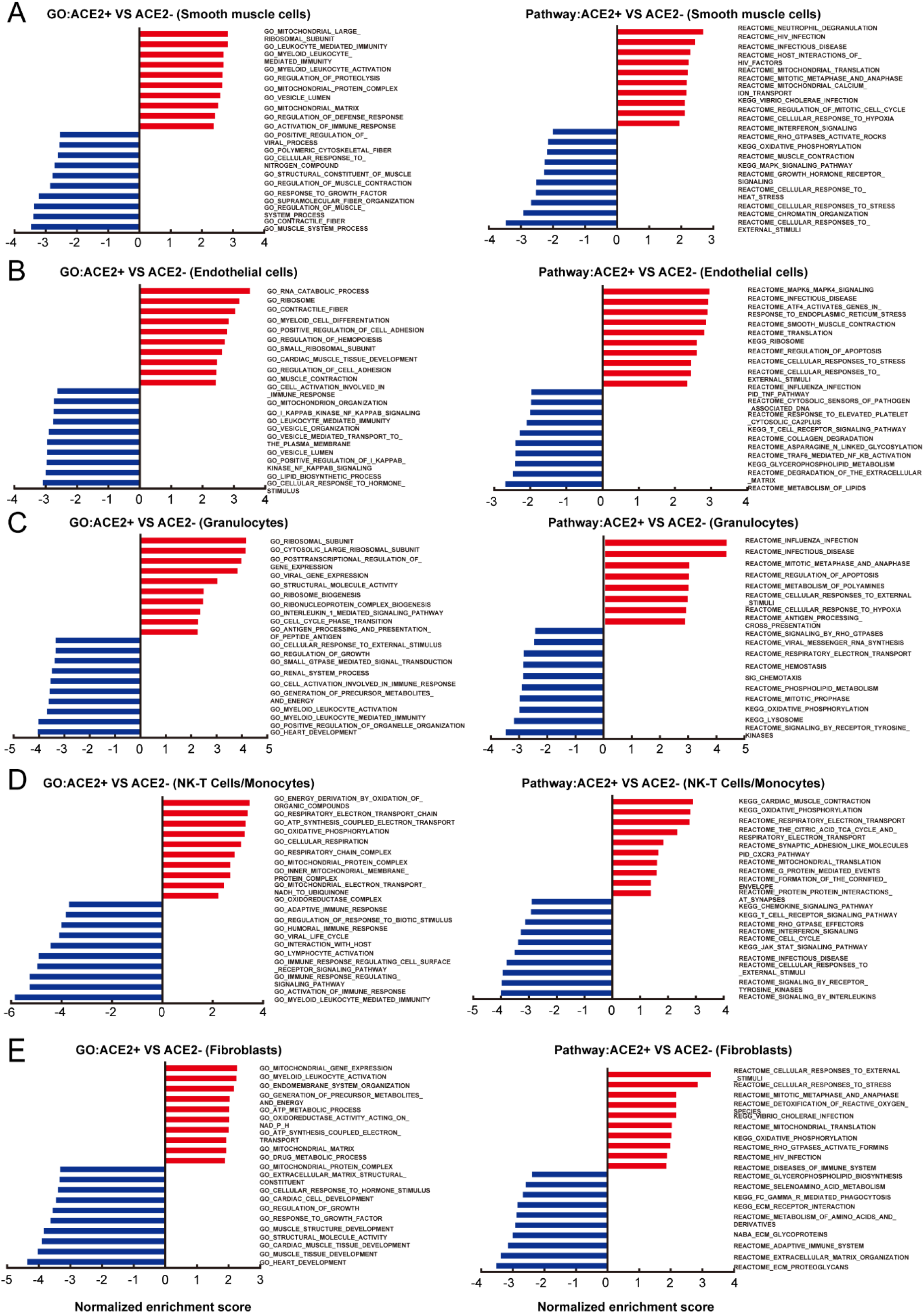
Enrichment of GO and biological pathway for ACE2+ compared with ACE2- in different subsets (related to Figure 5) **A-E**, GO and pathway analysis revealed the significant enrichment of biological pathway for ACE2+ compared with ACE2- cells in different subsets. **A**, Smooth muscle cells. **B**, endothelial cells. **C**, Granulocytes. **D**, NK-T cells/Monocytes. **E**, Fibroblasts.

## Notes

### Competing Interest Statement

The authors have declared no competing interest.

